# The therapeutic effect of intra-articular facet joint injection with normal saline as a comparator for chronic low back pain: a systematic review and meta-analysis

**DOI:** 10.1101/2021.01.27.21250595

**Authors:** Tanawin Nopsopon, Krit Pongpirul, Thanitsara Rittiphairoj, Irin Lertparinyaphorn, Areerat Suputtitada

## Abstract

**Background:** Intra-articular facet joint injection (FJI) has been increasingly used as a treatment for chronic low back pain (LBP). Choice of the substance has been based on clinical experience with unclear evidence on marginal effectiveness of active substance over normal saline as a placebo control. This systematic review investigates the comparative effectiveness between normal saline and active substances on patient-reported outcomes (PROs).

**Methods:** Systematic search was conducted in five databases: PubMed, Embase, Scopus, Web of Science, and CENTRAL for randomized controlled trials and observational studies of evaluating the PROs of FJI comparing active injected substances with normal saline as placebo in chronic LBP patients in the English language without publication date restriction. Quality assessment was performed using ROB2 and ROBINS-I. The meta-analysis was done using a random-effects model. Mean difference with 95% CIs of efficacy outcomes including pain, numbness, disability, quality of life were measured.

**Results:** Of 2,467 potential studies, three were included in the systematic review and meta-analysis (247 patients). Compared to other active substances, normal saline provided similar therapeutic effects on pain outcome within one hour (MD 2.43, 95% CI –11.61 to 16.50), at 1–1.5 months follow up (MD –0.63, 95% CI –7.97 to 6.72), and at 3 to 6 months (MD 1.90, 95% CI –16.03 to 19.83) as well as the quality of life at one and six months follow-up.

**Conclusions:** The short-term and long-term clinical improvements of intra-articular FJI using normal saline are comparable to the other active substances in LBP patients.

**PROSPERO:** registration number CRD42020216426

## INTRODUCTION

Chronic low back pain (LBP) is a common health problem of individuals at some point in adult life.^1^ Prevalence of chronic LBP had an increasing trend with aging and affected about one-fifth of the global population who were aged 20–59.^2^ Chronic LBP was the second leading cause of global work absence^3^ as well as the second most common chief complaint presented at physician office^4^ accounted for about 2.3% of all ambulatory visits, approximately 20.5 million visits each year^5^ and cost exceed US$100 billion per year in the United States.^6^

Non-operative treatments of chronic LBP were promoted as first-line treatment whereas a surgical option was considered only when non-operative treatment was not available or failed.^7^ More recent studies showed no superior long-term outcome of spinal fusion over non-operative treatment on pain and disability outcomes in patient with chronic LBP.^8 9^ For non-invasive treatment, most guidelines recommended education, exercise, manual therapy, multimodal rehabilitation, and oral medication including paracetamol, NSAIDs, and short-term opioids.^10 11^ Intra-articular FJI, which was considered the most invasive treatment of non-operative treatment, has been increasingly common^12 13^ despite no recommendation in most recent guidelines.^7 14 15^

The intra-articular FJI was developed while the authors attempted to find the pain pattern of facet syndrome used hypertonic saline and lidocaine as injected substances.^16^ Range of the injected substance options varied from the commonly used mixture of steroid and local anesthesia,^17^ steroids alone,^18^ and local anesthesia alone^19^ to more novel substances including ozone,^20^ autologous platelet-rich plasma (PRP),^21^ as well as hyaluronic acid (HA).^22^

Recent meta-analyses evaluated the therapeutic effects of intra-articular normal saline injection in knee osteoarthritis found significant pain reduction effects on both short-term and long-term follow up.^23 24^ A network meta-analysis that studied the effectiveness of various injected substances for intra-articular injection for hip osteoarthritis presented that no active substance was superior to normal saline for pain reduction.^25^ Nonetheless, these findings might not be generalized to intra-articular injection for lumbar facet joints. Although recent randomized controlled trials (RCTs) evaluating injected substances for intra-articular FJI usually compared novel treatment with active control,^21 22^ some RCTs compared between active substances and normal saline as a placebo control.^18^ Thus, evaluation regarding whether the efficacy and effectiveness demonstrated in normal saline intra-articular injection in the knee and the hip also showed therapeutic effects in the facet joint can be conducted. In this review, we performed a meta-analysis to evaluate whether intra-articular FJI with normal saline is similar to, if not better than, active substances for patient-reported outcomes (PROs).

## METHODS

### Study protocol

This study was conducted following the recommendations of the Preferred Reporting Items of Systematic Reviews and Meta-analyses (PRISMA) statement. We registered the systematic review with PROSPERO International Prospective Register of Ongoing Systematic Reviews (registration number: CRD42020216426).

### Search strategy

PubMed, Embase, Scopus, Web of Science, and CENTRAL were used to search for articles published in the English language up to 1 February 2020. The search strategy is presented in detail in the Supplement. Besides, the reference lists of included articles will be searched, as well as related citations from other journals via Google Scholar.

### Study selection

For this systematic review, we worked with an information specialist to design an appropriate search strategy to identify original peer-reviewed articles of RCTs and observational studies evaluating the PROs including pain, numbness, disability, quality of life, or complication outcomes of FJI comparing active injected substances with normal saline as placebo in patients with a diagnosis of chronic LBP. Article screening was done by two independent reviewers (TN and TR) for eligible studies. Discrepancies between the two reviewers were resolved by consensus.

### Data extraction

Data extraction was done by two independent reviewers (TN and IL) for published summary estimate data. Discrepancies between the two reviewers were resolved by consensus. We extracted the following data: (1) study characteristics (authors, year of publication, study type, journal name, contact information, country, and funding), (2) patient characteristics (sample size, age, age at onset, gender, comorbidities, method of diagnosis, inclusion and exclusion criteria, disease duration, location of back pain), (3) intervention (the type of injected active substances, dosage or regimen of injected substances, type of imaging guide, co-intervention), (4) comparators (volume of injected normal saline, approach technique, type of imaging guide, co-intervention), (5) outcomes (complete list of the names of all measured outcomes, unit of measurement, follow-up time point, missing data) as well as any other relevant information. All relevant text, tables, and figures were examined for data extraction. We contacted the authors of the study with incompletely reported data. If the trial authors did not respond within 14 days, we conducted analyses using the available data.

### Risk of bias assessment

The authors worked independently to assess the risk of bias in the included trials using the Cochrane Risk of Bias Tool 2.0 for RCT study.^26^ We assessed the randomization process, deviations from intended intervention, missing outcome data, measurement of the outcome, selection of the reported result. We Assigned each domain as a low risk of bias, some concerns, and a high risk of bias. For non-randomized trials and observational studies, we used the Risk of Bias In Non-randomized Studies of Interventions (ROBINS-I) to Investigate the confounding, selection of participants into the study, classification of interventions, deviations from intended interventions, missing data, measurement of outcomes, selection of the reported results.^27^ We assigned each domain as a low, moderate, serious, critical risk of bias, and no information. As mentioned above, we contacted the authors if there was insufficient information to assess. If the trial authors did not respond within 14 days, we conducted the assessment using available data. We resolved the disagreement through discussion, we present the risk of bias assessment in figure 2.

### Statistical analysis

The primary outcome was a visual analog scale (VAS) for the pain measurement tools. The outcomes measured were the mean difference (MD) in the reduction of VAS between before and after treatment with an associated 95% confidence interval (CI). Disability outcomes including Oswestry Disability Index (ODI) and Pain Disability Questionnaire (PDQ) were also retrieved with numbness outcome, quality of life outcome, and adverse events when reported. The results of the studies were included in the meta-analysis and presented in a forest plot, which also showed statistical powers, confidence intervals, and heterogeneity. We assessed clinical and methodological heterogeneity by examining participant characteristics, intervention regimen, type of intervention, follow-up period, outcomes, and study design. We then assessed statistical heterogeneity using the I^2^ and χ^2^ statistics. We regarded level of heterogeneity for I^2^ statistic as defined in chapter 9 of the Cochrane Handbook for Systematic Reviews of Interventions: 0–40% might not be important; 30–60% may represent moderate heterogeneity; 50–90% may represent substantial heterogeneity; 75–100% considerable heterogeneity. For missing standard deviation, we imputed the standard deviation as suggested in chapter 6 of the Cochrane Handbook for Systematic Reviews of Interventions referred to Furukawa methods.^28^ Sensitivity analysis was applied for an alternate method for imputing standard deviation if applicable. The random-effects meta-analysis by DerSimonian and Laird method was used as clinical, methodological, and statistical heterogeneity encountered. The meta-analysis was performed using Revman 5.3 (Cochrane Collaboration, Oxford, UK).

## RESULTS

### Study selection

The database search identified 2,467 potential records. After removing duplicates, 1,305 titles passed the initial Screen and 112 theme-related abstracts were selected for further full-text articles assessed for eligibility (figure 1). A total of 109 articles were excluded as the following: 47 wrong study designs, 24 wrong comparators, 23 non-peer-reviewed, six protocol, three duplicate, three non-English, two wrong intervention, and one wrong outcome. Only three studies were eligible for our inclusion criteria.

**Figure 1.**
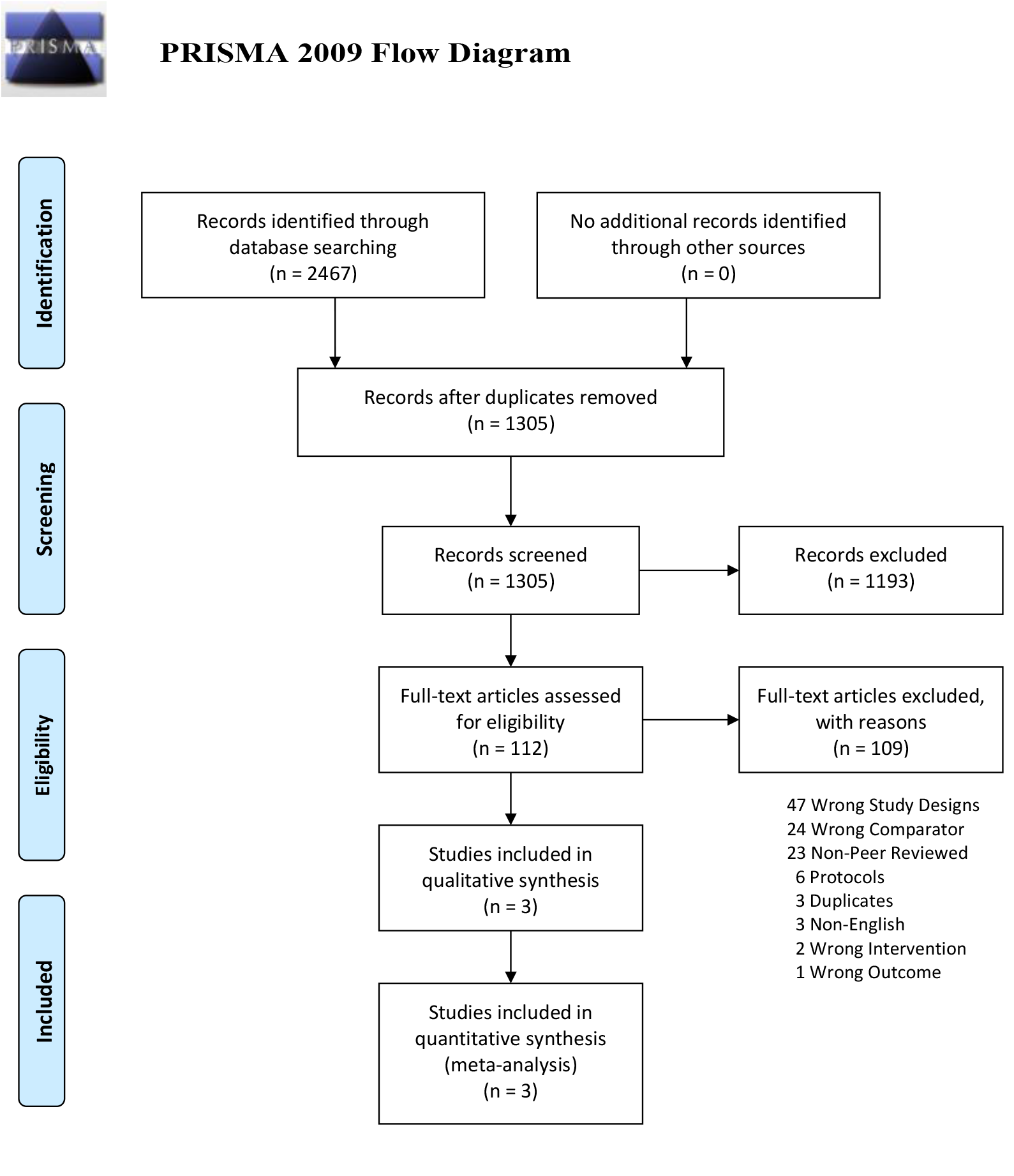
Flow chart diagram on how the studies were selected for analysis based on the Preferred Reporting Items for Systematic Reviews and Meta-analyses (PRISMA) guidelines.

### Study characteristics

The three included studies were published between 1989 and 1998 (table 1).^18 29 30^ There were three RCTs and zero non-randomized studies. The number of patients per study ranged from 70 to 97, with a total of 247 patients (137 of them were females, 55.5%). The mean age of included patient varied from 43-58 years. Two studies reported disease duration varied from a mean of 19.6 months, a median of 18 months in the intervention group, and a median of 24 months in the normal saline group. None of the included studies documented the co-morbidity and age at onset. Carette study was the only study reported the co-intervention involved 11 patients in corticosteroid and six patients in the placebo group.^18^ The included patients had a chronic LBP for over three to six months. The location of back pain also varied with L3/L4 to L5/S1. The follow-up period ranged from 0.5 hours to 6 months.

**Table 1.** Characteristics of the included studies

|  | Country | Interventions | Sample size | Disease duration (months) | Female ; n (%) | Age; mean (range) | Imaging technique | Patient reported outcomes |  |  |  |  |
| --- | --- | --- | --- | --- | --- | --- | --- | --- | --- | --- | --- | --- |
|  |  |  |  |  |  |  |  | Pain | Disability | Numbness | Quality of Life | Adverse events |
| Lilius | Finland | (1) 6 ml bupivacaine hydrochloride mixed with 2 ml methylprednisolone acetate<br>(2) 8 ml physiological saline | 70 | NR | 39 (56%) | 44 (19–64) | Fluoroscopy | VAS | Objective disability score (Only overall result) | NR | NR | 7 Overall (5 men, 2 women) |
| Carette | Canada | (1) 1 ml methyl prednisolone acetate mixed with 1 ml of isotonic saline<br>(2) 2 ml isotonic saline | 97 | (1) median 18 months<br>(2) median 24 months | 44 (45%) | 43 | Fluoroscopy | VAS | NR | NR | SIPs | No adverse events occurred |
| Revel | France | (1) 1 ml 2% lidocaine<br>(2) 1 ml normal saline | 80 | Overall mean 19.6 months | 54 (68%) | 58 (34–87) | Fluoroscopy | VAS | NR | NR | NR | NR |

NR, not reported; VAS, visual analog scale.

None of the three RCTs had the same type of active substances in the intervention arm. One study had a mixture of corticosteroid and local anesthesia as the intervention.^29^ One study used corticosteroid alone while the other one used local anesthesia alone.^18 30^ All three trials used fluoroscopy-guided for FJI.

### Quality Assessment

For the risk of bias assessment, the three randomized controlled trials included in this study had adequate randomization process in one trial, ensured deviations from intended interventions in two trials, ensured missing outcome data in two trials, adequate measurement of the outcome in three trials, and ensured the selection of the reported result in one trial.^18 29 30^ Only a high risk of bias in the selection of the reported result presented in one trial. A summary of the risk of bias assessment of randomized controlled trials which were at low, some concerns, and high risk for each risk of bias domain was presented in figure 2.

**Figure 2.**
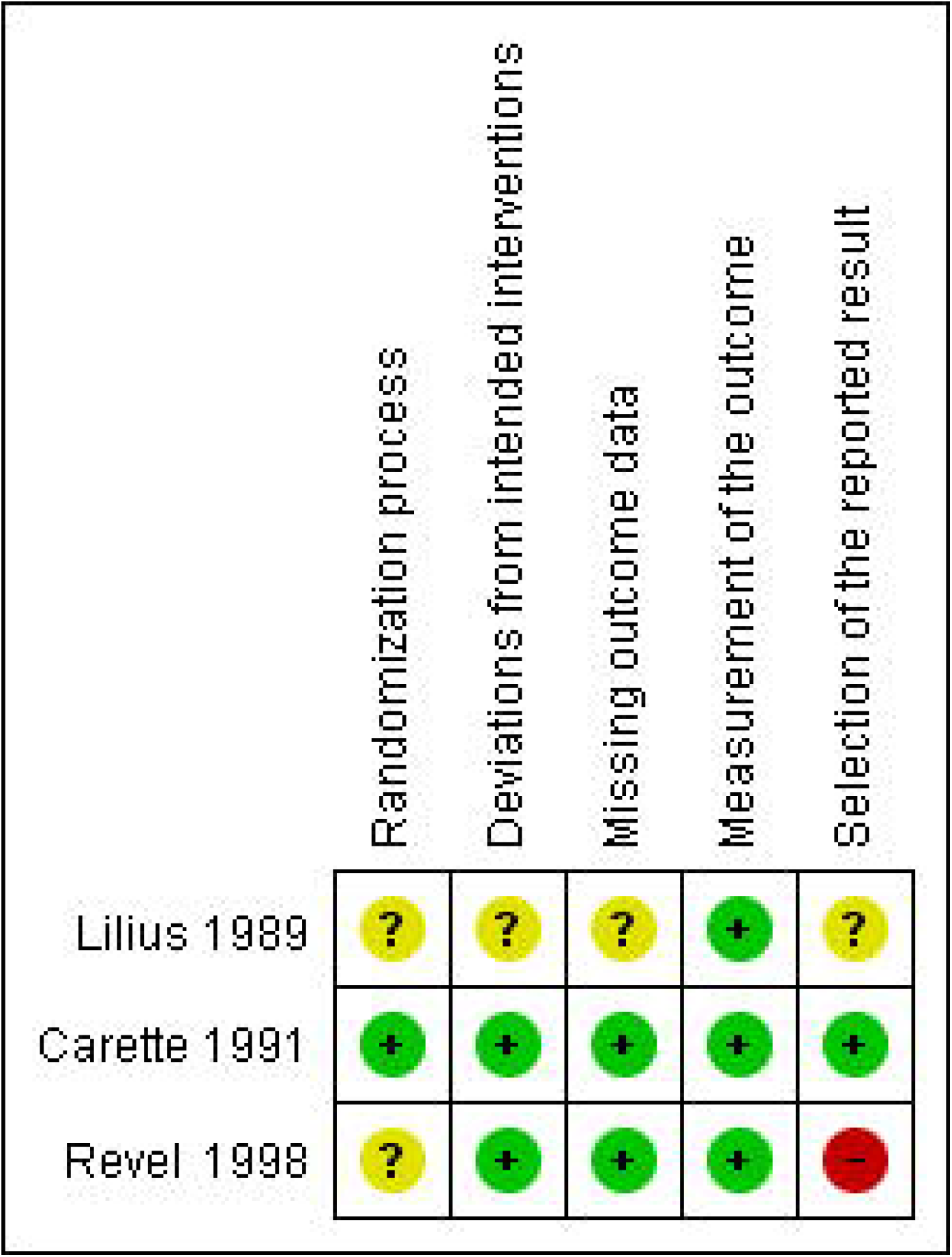
Risk of biases of included randomized controlled trials.

### Qualitative analysis

The primary outcome assessed by visual analog pain scale (VAS) has been examined in all included studies, consisted of three randomized controlled trials with a total number of 247 patients. Only Lilius study reported disability outcome using a combined score proposed by the authors on a whole study group, but it was not reported disability outcome for each intervention group.^29^ Carette study reported no difference between corticosteroid and normal saline FJI in overall quality of life outcome using Sickness Impact Profile (SIP) score which consisted of physical and psychosocial dimensions both on one and six months follow up while only significant difference was reported on the physical dimension on six months follow up with a favorable on corticosteroid injection (mean difference (MD) –3.5, 95% CI –6.2 to –0.9).^18^ Two studies reported adverse events.^18 29^ Lilius study reported seven overall adverse events, five in men and two in women,^29^ while Carette study reported no major adverse event related to intra-articular FJI.^18^ Unfortunately, only one study reported the eligible result in each outcome, therefore the meta-analysis could not be conducted for the disability, quality of life, and adverse event outcomes. None of the three included trials investigated numbness outcome.

Lilius study demonstrated a significant reduction of subjective pain for all participants at all follow-up points (one hour, two weeks, six weeks, and three months), but there was no significant difference in subjective visual pain scale reduction for intra-articular injection with a mixture of local anesthesia and steroid compared with the intra-articular normal saline injection for facet joint L3/4 to L5/S1 for both short-term and long-term response range from one hour to three months after injection.^29^

Carette study demonstrated no difference between corticosteroid and normal saline injection in pain measurement outcome using McGill pain questionnaire ^31^ both one and six months follow up. The author also investigated on self-rated pain assessment reported with very marked or marked improvement which was not a significant difference on one month follow up, but patients treated with corticosteroid FJI reported a favorable marked improvement on six months follow-up (MD 31%, 95% CI 14 to 48%).^18^

Revel study concluded that FJI with local anesthesia had a significant benefit in the change of pain score over FJI with normal saline (p=0.01) in a specific patient group who met five or more of the seven clinical characteristics proposed by the author consisted of 1) age greater than 65 years, 2) no pain exacerbation by cough, 3) no pain exacerbation by forward-flexion, 4) no pain exacerbation when rising from flexion, 5) no pain exacerbation by hyperextension, 6) no pain exacerbation by extension-rotation, and 7) pain well relieved by recumbent. For the patient who did not meet five items of the criteria, FJI with normal saline showed a trend of better pain improvement, although not significantly.^30^

### Quantitative analysis

All three studies, which reported the pain outcome assessed by visual analog pain scale, were included in the meta-analysis.^18 29 30^ Revel study showed a significantly favorable outcome with VAS reduction after FJI with local anesthesia within one hour follow up,^30^ whereas Lilius study showed a trend of the benefit in favor the treatment with normal saline injection; however, it was not significantly.^29^ The pooled effect of two randomized controlled trials showed no difference in pain reduction within one hour follow up between FJI with active substances and normal saline (MD 2.43, 95% CI –11.61 to 16.50) (figure 3).

**Figure 3.**
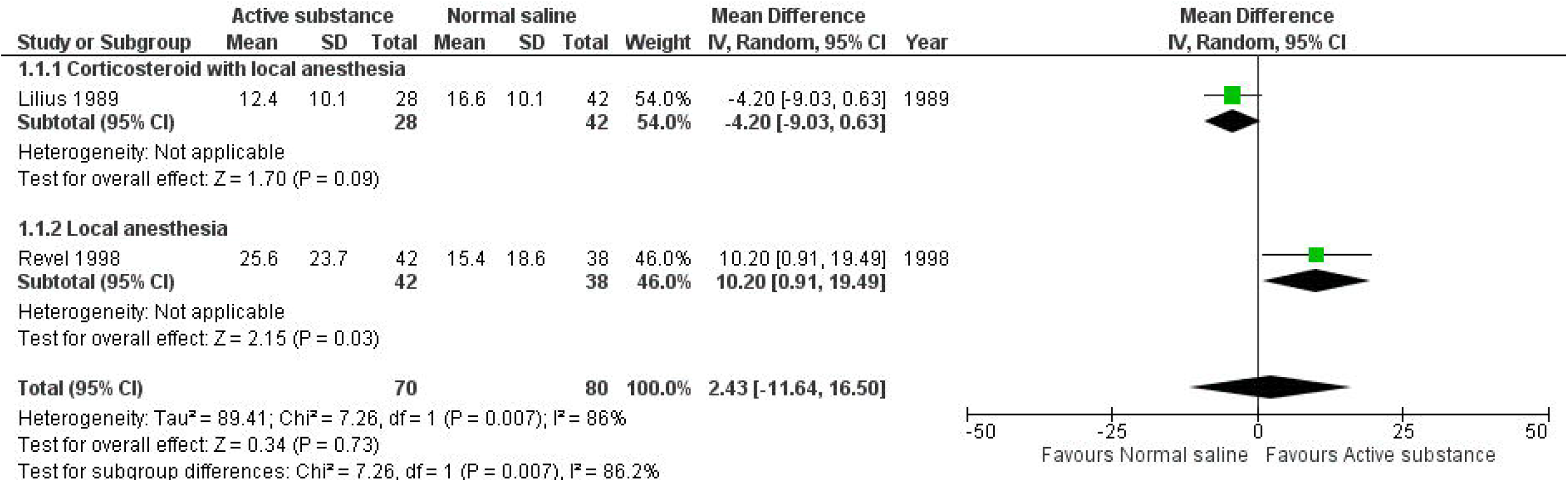
Forest plot showing the effect of active substance versus normal saline on pain reduction within one hour.

Lilius and Carette studies reported no difference in pain diminishing between active substances and normal saline FJI as placebo with 1–1.5 months follow up.^18 29^ The meta-analysis of the two studies showed a similar result; the mean difference was –0.63 (95% CI –7.97 to 6.72) (figure 4).

**Figure 4.**
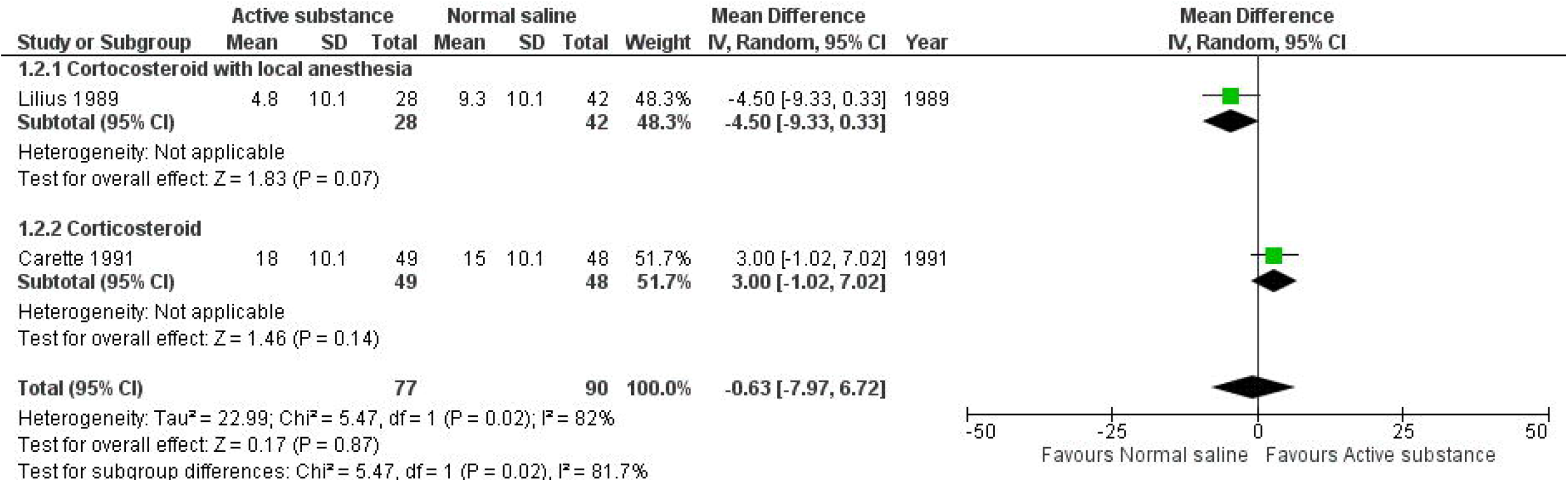
Forest plot showing the effect of active substance versus normal saline on pain reduction at 1–1.5 months follow-up visit.

Two studies reported long-term follow-up in pain scores ranged from 3 to 6 months. Lilius study reported a significantly favorable benefit of normal saline over active substance on long-term pain reduction^29^ while Carette study reported a significantly favorable benefit of corticosteroid over placebo intra-articular FJI on long-term pain reduction.^18^ The pooled effects of long-term pain reduction after FJI showed no difference between injected with active substances and injected with normal saline as placebo (MD 1.90, 95% CI –16.03 to 19.83) (figure 5).

**Figure 5.**
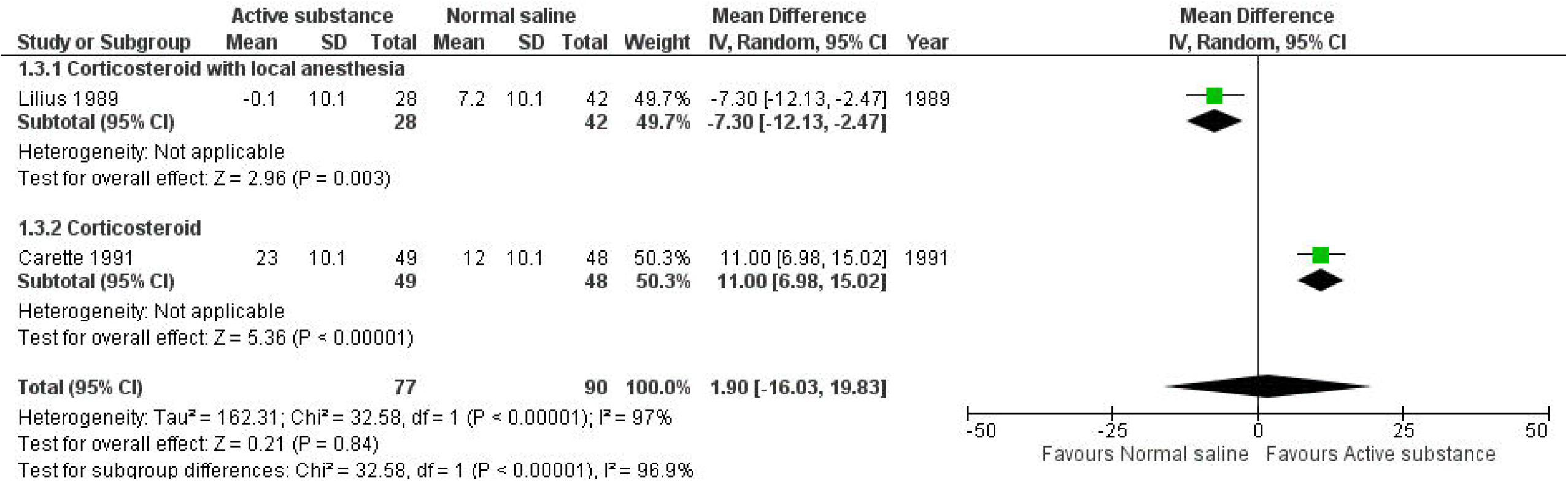
Forest plot showing the effect of active substance versus normal saline on pain reduction at 3–6 months follow-up visit.

Sensitivity analysis was performed with an alternative method of standard deviation imputation using standard deviation from the included study in this meta-analysis instead of from the previous meta-analysis. The pooled effect of pain reduction within one hour, with 1–1.5 months follow up, and with long-term follow up was similar to the main imputation method (online supplementary file).

## DISCUSSION

Our meta-analysis suggests that the treatment with normal saline intra-articular FJI as placebo provided similar effectiveness to intra-articular FJI with active substance for patient-reported pain outcome measured by visual analog pain scales at all studied time point ranged from 0.5 hours to six months after FJI.

It is hard to conclude from the trials because of the heterogeneity presented in the age of population targets, type of active substance, location of back pain, follow-up time, and outcome measurements. The findings suggested no difference in the pain reduction benefit between FJI with normal saline and with active substances for chronic LBP patients; however, they needed more robustness to provide a high level of evidence.

Lilius study showed significant pain reduction for both saline and steroid with local anesthesia FJIs while no significant differences between intra-articular saline and intra-articular steroid with local anesthesia injection on subjective pain scale at all follow-up ranged from one hour to three months. One-fourth of subjects experienced pain reduction benefit up to three months after FJI and overall disability score was significantly improved regardless of the injected substance. The therapeutic effect of saline was unexpected, and only suggestion on the psychosocial point of view and self-regression had been provided without clear evidence and explanation.^29^

Carette study demonstrated a similar LBP reduction effect of corticosteroid FJI to normal saline FJI at one- and six-months follow-up evaluated with visual analog pain scale and McGill pain questionnaire. The authors conclude that FJI with corticosteroid provided little benefit to patients with chronic LBP with the belief that normal saline injection was a true placebo. Moreover, this study was the only one in meta-analysis assessed the quality of life outcome using Sickness Impact Profile score with the only favorable effect of steroid could be observed for physical dimension at six months follow up, but not for psychosocial dimension.^18^

Revel study explored the characteristics of chronic LBP patients which were significant predictors of a favorable pain reduction of local anesthesia intra-articular FJI compared to saline FJI which consisted of five characteristics of present back pain. Normal saline was considered as a true placebo and the therapeutic effect of saline was suspected for inadequate diagnostic criteria which resulted in a false-positive selection of chronic LBP patients who would potentially have benefited from FJI.^30^

High-quality systematic review and meta-analysis had been conducted to identify injection therapy for subacute and chronic LBP; only one study compared FJI with the active substance and with placebo^18^ had been identified and included in a meta-analysis of pain outcome with self-compared between short-term and long-term therapeutic effect which resulted in no significant difference.^32^ More recent systematic review attempt to compared efficacy of saline, local anesthesia, and steroids in FJI also identified the same study^18^ without conducting meta- analysis.^33^ Thus, our study is the first meta-analysis conducted with more than one included study.

Two meta-analyses focused on the efficacy of intra-articular normal saline injection for knee osteoarthritis demonstrated therapeutic effects of normal saline injection on pain^23 24^ and functional outcome;^24^ however, the meta-analyses conducted comparing pre-injection and post-injection, not between injected substance. Network meta-analysis evaluated the effectiveness of various injected substance for intra-articular injection in patients with hip osteoarthritis showed that intra-articular hip saline injection had similar effects to all other active substances on pain and functional outcome.^25^ These studies provided strong evidence for the potential therapeutic intra-articular saline injection which were concordance with our findings, which cast the doubt on the appropriateness of using normal saline intra-articular as true placebo and the result interpretation.

The paradox for practices of intra-articular FJI had occurred. No early trials could provide any significant benefit of the active substance over normal saline as the placebo,^18 29 30^ resulted in lack of supporting evidence to recommended the use of intra-articular FJI in guidelines;^7 14 15^ however, the trend of utilization intra-articular FJI in real-world practices was increasing.^13^ Another paradox occurred for the choice of injected substance for intra-articular FJI when more recent trials chose a combination of corticosteroid and local anesthesia^21^ or corticosteroid alone^22^ as a comparator of a novel injected substance instead of normal saline as a true placebo comparator despite no evidence of a superior benefit of local anesthesia or corticosteroid over the normal saline intra-articular FJI. The result interpretation would have a serious problem, especially when no significant difference was observed: a novel substance has a similar effect to corticosteroid or local anesthesia, or a combination of both which have similar effect to normal saline, a true placebo. In the past, many authors considered that normal saline would actually be a true placebo and concluded that there was no therapeutic benefit of intra-articular FJI on LBP.^18 30^ Results from our study might be a missing piece of jigsaw that there was plausible evidence demonstrated that normal saline was not a true placebo, thus intra-articular FJI might have benefits for chronic LBP all along.

Normal saline therapeutic effect on pain reduction had been demonstrated in meta-analyses on osteoarthritis of knee^23 24^ and hip.^25^ However, the explanation of the effect had been rarely studied and mostly based on hypothesis. One hypothesis is the dilution of inflammatory mediators which resulted in pain relief.^34^ A study attempted to explore the potential of other mechanisms including the osmolality effect and sodium concentration, but no sufficient evidence to support the hypothesis.^35^ For facet joint, the first saline injection which resulted in pain relief was hypertonic saline.^36^ Caterini study found that facet joint pain might originate from excessive facet joint fluid,^37^ thus this could be an explanation on how hypertonic saline could relieve facet joint pain, but not for normal saline. The osmolality effect might come into account since facet joint pain could originate from various causes, there might be some type of facet joint pain that the patients had a normal volume of facet joint fluid but had imbalance osmolality. This is merely one of the hypotheses since no available study on this question.

There were several limitations in this meta-analysis. First, the presence of heterogeneous characteristics of the patients, type of injected substance, and timing of outcome assessment led to difficulty for the conclusion of the data. Second, some trials did not report the standard deviation needed for the meta-analysis, thus techniques of the imputation of missing standard deviation had been used which might not reflect the real variation in outcome for the study. Third, there was a small number of studies focused on injected substance for intra-articular FJI which led to only a few studies has been included in the meta-analysis and some PROs had insufficient outcome to conduct a meta-analysis. Fourth, there was surprisingly no new trial compared active substance and normal saline for twenty years. The evidence might not be completely comparable to present practices for intra-articular JFI; however, this study provided the most up-to-date pieces of evidence to shed light on the therapeutic effects of intra-articular normal saline FJI.

In conclusion, this systematic review and meta-analysis suggests that the short-term and long-term clinical improvements of intra-articular FJI using normal saline are comparable to the other active substances in LBP patients.

## Supporting information

Supplemental Material [PRISMA Checklist]

## Data Availability

Data are available on reasonable request.

## Contributors

TN took part in design of this project, acquisition of data, analysis and interpretation of data, and writing the manuscript. KP took part in conception and design of this project, interpretation of data, and writing the manuscript. TR took part in acquisition of data, interpretation of data, and writing the manuscript. IL took part in interpretation of data and writing the manuscript. AS took part in conception and design of this project, and writing the manuscript.

## Funding

The authors have not declared a specific grant for this research from any funding agency in the public, commercial or not-for-profit sectors.

## Competing interests

None declared.

## Patient consent for publication

Not required.

## Data availability statement

Data are available on reasonable request.

## Notes

### Competing Interest Statement

The authors have declared no competing interest.

### Clinical Trial

PROSPERO registration number CRD42020216426

