## Supplemental Material [Figures and Appendix] for "The therapeutic effect of intra-articular facet joint injection with normal saline as a comparator for chronic low back pain: a systematic review and meta-analysis"

**Supplementary Materials**

1. **Supplementary Figures**

   **Supplementary Figure S1. Percentage of risk of biases of included randomized controlled trials.**

   **Supplementary Figure S2. Forest plots: Sensitivity analysis with alternative method for standard deviation imputation.**

2. **Appendix**

   **Appendix 1. Full search strategy**

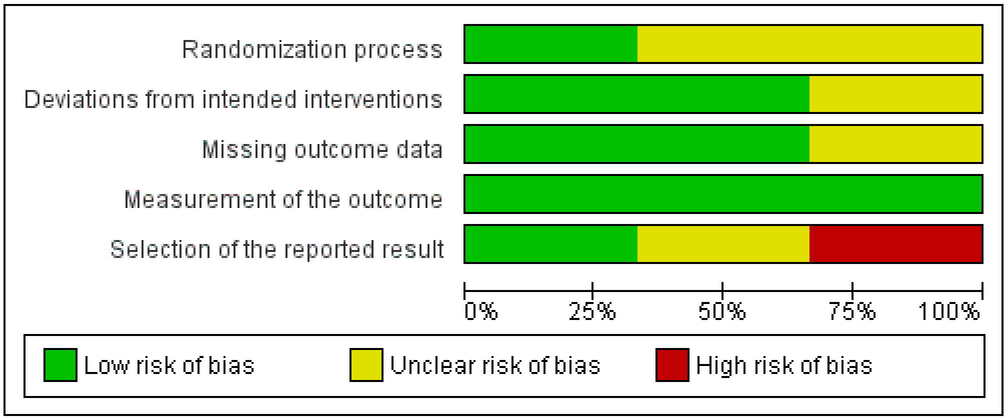


**Figure S1** Percentage of risk of biases of included randomized controlled trials.

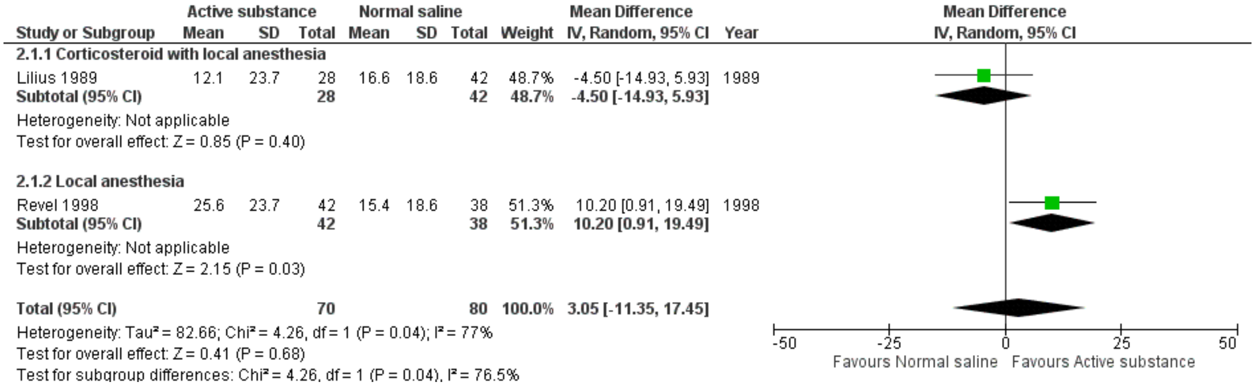


**Figure S2A** Forest plot showing the effect of active substance versus normal saline on pain reduction within one hour with alternative method for standard deviation imputation.

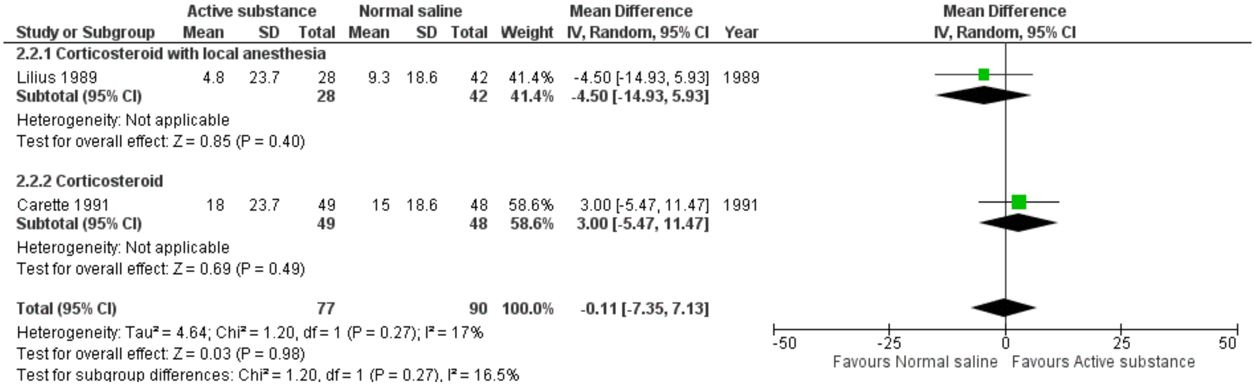


**Figure S2B** Forest plot showing the effect of active substance versus normal saline on pain reduction at 1–1.5 months follow-up visit with alternative method for standard deviation imputation.

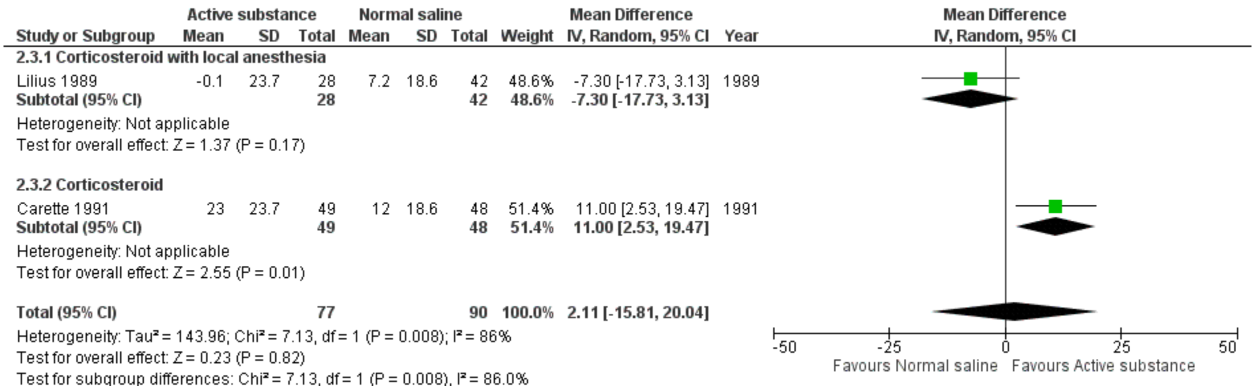


**Figure S2C** Forest plot showing the effect of active substance versus normal saline on pain reduction at 3–6 months follow-up visit with alternative method for standard deviation imputation.

**Appendix 1A** Pubmed search strategy

#1 "zygapophyseal joint"[MeSH Terms]
#2 zygapophyseal[tiab]
#3 zygapophysial[tiab]
#4 facet[tiab]
#5 (#2 OR #3 OR #4)
#6 joint[tiab]
#7 joints[tiab]
#8 (#6 OR #7)
#9 (#5 AND #8)
#10 "zygapophyseal joint"[tiab]
#11 "zygapophyseal joints"[tiab]
#12 "zygapophysial joint"[tiab]
#13 "zygapophysial joints"[tiab]
#14 "facet joint"[tiab]
#15 "facet joints"[tiab]
#16 (#1 OR #9 OR #10 OR #11 OR #12 OR #13 OR #14 OR #15)
#17 "injections, spinal"[MeSH Terms]
#18 injection[tiab]
#19 injections[tiab]
#20 intervention[tiab]
#21 interventions[tiab]
#22 (#17 OR #18 OR #19 OR #20 OR #21)
#23 (#16 AND #22)
#24 "facet joint injection"[tiab]
#25 fji[tiab]
#26 facet[tiab]
#27 block[tiab]
#28 blocks[tiab]
#29 (#27 OR #28)
#30 (#26 AND #29)
#31 "facet block"[tiab]
#32 "facet blocks"[tiab]
#33 (#23 OR #24 OR #25 OR #30 OR #31 OR #32)
#34 "back pain"[MeSH Terms]

#35 back[tiab]
#36 vertebrogenic[tiab]
#37 spine[tiab]
#38 spinal[tiab]
#39 lumbar[tiab]
#40 thoracic[tiab]
#41 (#35 OR #36 OR #37 OR #38 OR #39 OR #40)
#42 pain[tiab]
#43 pains[tiab]
#44 ache[tiab]
#45 aches[tiab]
#46 aching[tiab]
#47 (#42 OR #43 OR #44 OR #45 OR #46)
#48 (#41 AND #47)
#49 "back pain"[tiab]
#50 "back pains"[tiab]
#51 "back ache"[tiab]
#52 backache[tiab]
#53 "back aches"[tiab]
#54 backaches[tiab]
#55 "vertebrogenic pain"[tiab]
#56 "vertebrogenic pains"[tiab]
#57 "vertebrogenic pain syndrome"[tiab]
#58 "vertebrogenic pain syndromes"[tiab]
#59 dorsalgia[tiab]
#60 dorsalgias[tiab]
#61 "low back pain"[MeSH Terms]
#62 "low back pain"[tiab]
#63 lumbago[tiab]
#64 "sciatica"[MeSH Terms]
#65 sciatica[tiab]
#66 sciatic[tiab]
#67 neuralgia*[tiab]
#68 (#66 AND #67)
#69 "sciatic neuralgia"[tiab]
#70 "sciatic neuralgias"[tiab]
#71 claudication[tiab]
#72 "pain, referred"[MeSH Terms]
#73 refer[tiab]
#74 referred[tiab]
#75 hip[tiab]
#76 knee[tiab]
#77 leg[tiab]
#78 (#73 OR #74 OR #75 OR #76 OR #77)
#79 (#47 AND #78)
#80 "refer pain"[tiab]
#81"referred pain"[tiab]
#82 "hip pain"[tiab]
#83 "hip pains"[tiab]
#84 "knee pain"[tiab]
#85 "knee pains"[tiab]
#86 "leg pain"[tiab]

#87 "leg pains"[tiab]
#88 "paresthesia"[MeSH Terms]
#89 paresthesia[tiab]
#90 paresthesias[tiab]
#91 numbness[tiab]
#92 (#48 OR #49 OR #50 OR #51 OR #52 OR #53 OR #54 OR #55 OR #56 OR #57 OR #58 OR #59 OR #60 OR #61 OR #62 OR #63 OR #64 OR #65 OR #68 OR #69 OR #70 OR #71 OR #72 OR #79 OR #80 OR #81 OR #82 OR #83 OR #84 OR #85 OR #86 OR #87 OR #88 OR #89 OR #90 OR #91)
#93 ae[sh]
#94 co[sh]
#95 de[sh]
#96 safe[tiab]
#97 safety[tiab]
#98 "side effect"[tiab]
#99 "side effects"[tiab]
#100 "undesirable effect"[tiab]
#101 "undesirable effects"[tiab]
#102 "treatment emergent"[tiab]
#103 "treatment emergency"[tiab]
#104 tolerability[tiab]
#105 tolerance[tiab]
#106 tolerate[tiab]
#107 toxicity[tiab]
#108 toxic[tiab]
#109 harm[tiab]
#110 harms[tiab]
#111 complication*[tiab]
#112 risk[tiab]
#113 risks[tiab]
#114 "unintended event"[tiab]
#115 "unintended events"[tiab]
#116 "unintended effect"[tiab]
#117 "unintended effects"[tiab]
#118 "adverse event"[tiab]
#119 "adverse events"[tiab]
#120 "adverse effect"[tiab]
#121 "adverse effects"[tiab]
#122 "adverse reaction"[tiab]
#123 "adverse reactions"[tiab]
#124 "adverse outcome"[tiab]
#125 "adverse outcomes"[tiab]
#126 (#93 OR #94 OR #95 OR #96 OR #97 OR #98 OR #99 OR #100 OR #101 OR #102 OR #103 OR #104 OR #105 OR #106 OR #107 OR #108 OR #109 OR #110 OR #111 OR #112 OR #113 OR #114 OR #115 OR #116 OR #117 OR #118 OR #119 OR #120 OR #121 OR #122 OR #123 OR #124 OR #125)
#127 "spinal stenosis"[MeSH Terms]
#128 "spondylosis"[MeSH Terms]
#129 "synovial cyst"[MeSH Terms]
#130 back[tiab]
#131 vertebral[tiab]
#132 spine[tiab]
#133 spinal[tiab]
#134 lumbar[tiab]

#135 thoracic[tiab]
#136 facet[tiab]
#137 synovial[tiab]
#138 (#130 OR #131 OR #132 OR #133 OR #134 OR #135 OR #136 OR #137)
#139 stenosis[tiab]
#140 stenoses[tiab]
#141 spondylosis[tiab]
#142 syndrome[tiab]
#143 syndromes[tiab]
#144 arthropathy[tiab]
#145 arthritis[tiab]
#146 fracture[tiab]
#147 fractures[tiab]
#148 cyst[tiab]
#149 cysts[tiab]
#150 (#139 OR #140 OR #141 OR #142 OR #143 OR #144 OR #145 OR #146 OR #147 OR #148 OR #149)
#151 (#138 AND #150)
#152 "vertebral canal stenosis"[tiab]
#153 "spinal stenosis"[tiab]
#154 "spinal stenoses"[tiab]
#155 "thoracic spondylosis"[tiab]
#156 "lumbar spondylosis"[tiab]
#157 "facet syndrome"[tiab]
#158 "facet joint syndrome"[tiab]
#159 "facet arthropathy"[tiab]
#160 "facet arthritis"[tiab]
#161 "vertebral compression fracture"[tiab]
#162 "spinal cyst"[tiab]
#163 "facet cyst"[tiab]
#164 "synovial cyst"[tiab]
#165 "tarlov cysts"[MeSH Terms]
#166 "tarlov cyst"[tiab]
#167 "tarlov cysts"[tiab]
#168 "spondylolysis"[MeSH Terms]
#169 spondylolysis[tiab]
#170 spondylolyses[tiab]
#171 "spondylolisthesis"[MeSH Terms]
#172 spondylolisthesis[tiab]
#173 spondylolistheses[tiab]
#174 "myofascial pain syndromes"[MeSH Terms]
#175 "myofascial pain syndrome"[tiab]
#176 "myofascial pain syndromes"[tiab]
#177 (#127 OR #128 OR #129 OR #151 OR #152 OR #153 OR #154 OR #155 OR #156 OR #157 OR #158 OR #159 OR #160 OR #161 OR #162 OR #163 OR #164 OR #165 OR #166 OR #167 OR #168 OR #169 OR #170 OR #171 OR #172 OR #173 OR #174 OR #175 OR #176)
#178 (#92 OR #126)
#179 (#33 AND #177 AND #178)
#180 animals[MeSH Terms]
#181 humans[MeSH Terms]
#182 (#180 NOT #181)
#183 (#179 NOT #182)

#184 english[lang]
#185 (#183 AND #184)

**Appendix 1B** Embase search strategy

#1 'zygaphophyseal joint'/exp
#2 'zygapophyseal':ti,ab
#3 'zygapophysial':ti,ab
#4 'facet':ti,ab
#5 (#2 OR #3 OR #4)
#6 'joint*':ti,ab
#7 (#5 AND #6)
#8 'zygapophyseal joint':ti,ab
#9 'zygapophyseal joints':ti,ab
#10 'zygapophysial joint':ti,ab
#11 'zygapophysial joints':ti,ab
#12 'facet joint':ti,ab
#13 'facet joints':ti,ab
#14 (#1 OR #7 OR #8 OR #9 OR #10 OR #11 OR #12 OR #13)
#15 'intraspinal drug administration'/exp
#16 'injection*':ti,ab
#17 'intervention*':ti,ab
#18 (#15 OR #16 OR #17)
#19 (#14 AND #18)
#20 'facet joint injection':ti,ab
#21 'fji':ti,ab
#22 'facet':ti,ab
#23 'block*':ti,ab
#24 (#22 AND #23)
#25 'facet block':ti,ab
#26 'facet blocks':ti,ab
#27 (#19 OR #20 OR #21 OR #24 OR #25 OR #26)
#28 'backache'/exp
#29 'back':ti,ab
#30 'vertebrogenic':ti,ab
#31 'spine':ti,ab
#32 'spinal':ti,ab
#33 'lumbar':ti,ab
#34 'thoracic':ti,ab
#35 (#29 OR #30 OR #31 OR #32 OR #33 OR #34)
#36 'pain*':ti,ab
#37 'ache*':ti,ab
#38 'aching':ti,ab
#39 (#36 OR #37 OR #38)
#40 (#35 AND #39)
#41 'back pain':ti,ab
#42 'back pains':ti,ab
#43 'back ache':ti,ab
#44 'backache':ti,ab
#45 'back aches':ti,ab
#46 'backaches':ti,ab

#47 'vertebrogenic pain':ti,ab
#48 'vertebrogenic pains':ti,ab
#49 'vertebrogenic pain syndrome':ti,ab
#50 'vertebrogenic pain syndromes':ti,ab
#51 'dorsalgia':ti,ab
#52 'dorsalgias':ti,ab
#53 'low back pain'/exp
#54 'low back pain':ti,ab
#55 'lumbago':ti,ab
#56 'sciatica'/exp
#57 'sciatica':ti,ab
#58 'sciatic':ti,ab
#59 'neuralgia*':ti,ab
#60 (#58 AND #59)
#61 'sciatic neuralgia':ti,ab
#62 'sciatic neuralgias':ti,ab
#63 'claudication':ti,ab
#64 'referred pain'/exp
#65 'refer':ti,ab
#66 'referred':ti,ab
#67 'hip':ti,ab
#68 'knee':ti,ab
#69 'leg':ti,ab
#70 (#65 OR #66 OR #67 OR #68 OR #69)
#71 (#39 AND #70)
#72 'refer pain':ti,ab
#73 'referred pain':ti,ab
#74 hip pain':ti,ab
#75 'hip pains':ti,ab
#76 'knee pain':ti,ab
#77 'knee pains':ti,ab
#78 'leg pain':ti,ab
#79 'leg pains':ti,ab
#80 'paresthesia'/exp
#81 'paresthesia':ti,ab
#82 'paresthesias':ti,ab
#83 'numbness':ti,ab
#84 (#28 OR #40 OR #41 OR #42 OR #43 OR #44 OR #45 OR #46 OR #47 OR #48 OR #49 OR #50 OR #51 OR #52 OR #53 OR #54 OR #55 OR #56 OR #57 OR #60 OR #61 OR #62 OR #63 OR #64 OR #71 OR #72 OR #73 OR #74 OR #75 OR #76 OR #77 OR #78 OR #79 OR #80 OR #81 OR #82 OR #83)
#84 'adverse drug reaction'/lnk
#85 'adverse device effect'/lnk
#86 'complication'/lnk
#87 'side effect'/lnk
#88 'safe':ti,ab
#89 'safety':ti,ab
#90 'side effect':ti,ab
#91 'side effects':ti,ab
#92 'undesirable effect':ti,ab
#93 'undesirable effects':ti,ab
#94 'treatment emergent':ti,ab
#95 'treatment emergency':ti,ab

#96 'tolerability':ti,ab
#97 'tolerance':ti,ab
#98 'tolerate':ti,ab
#99 'toxicity':ti,ab
#100 'toxic':ti,ab
#101 'harm*':ti,ab
#102 'complication*':ti,ab
#103 'risk*':ti,ab
#104 'unintended event':ti,ab
#105 'unintended events':ti,ab
#106 'unintended effect':ti,ab
#107 'unintended effects':ti,ab
#108 'adverse event':ti,ab
#109 'adverse events':ti,ab
#110 'adverse effect':ti,ab
#111 'adverse effects':ti,ab
#112 'adverse reaction':ti,ab
#113 'adverse reactions':ti,ab
#114 'adverse outcome':ti,ab
#115 'adverse outcomes':ti,ab
#116 (#84 OR #85 OR #86 OR #87 OR #88 OR #89 OR #90 OR #91 OR #92 OR #93 OR #94 OR #95 OR #96 OR #97 OR #98 OR #99 OR #100 OR #101 OR #102 OR #103 OR #104 OR #105 OR #106 OR #107 OR #108 OR #109 OR #110 OR #111 OR #112 OR #113 OR #114 OR #115)
#116 'vertebral canal stenosis'/exp
#117 'spondylosis'/exp
#118 'synovial cyst'/exp
#119 'back':ti,ab
#120 'vertebral':ti,ab
#121 'spine':ti,ab
#122 'spinal':ti,ab
#123 'lumbar':ti,ab
#124 'thoracic':ti,ab
#125 'facet':ti,ab
#126 'synovial':ti,ab
#127 (#119 OR #120 OR #121 OR #122 OR #123 OR #124 OR #125 OR #126)
#128 'stenosis':ti,ab
#129 'stenoses':ti,ab
#130 'spondylosis':ti,ab
#131 'syndrome*':ti,ab
#132 'arthropathy':ti,ab
#133 'arthritis':ti,ab
#134 'fracture*':ti,ab
#135 'cyst*':ti,ab
#136 (#128 OR #129 OR #130 OR #131 OR #132 OR #133 OR #134 OR #135)
#137 (#127 AND #136)
#138 'vertebral canal stenosis':ti,ab
#139 'spinal stenosis':ti,ab
#140 'spinal stenoses':ti,ab
#141 'thoracic spondylosis':ti,ab
#142 'lumbar spondylosis':ti,ab
#143 'facet syndrome':ti,ab
#144 'facet joint syndrome':ti,ab

#145 'facet arthropathy':ti,ab
#146 'facet arthritis':ti,ab
#147 'vertebral compression fracture':ti,ab
#148 'spinal cyst':ti,ab
#149 'facet cyst':ti,ab
#150 'synovial cyst':ti,ab
#151 'tarlov cyst'/exp
#152 'tarlov cyst':ti,ab
#153 'tarlov cysts':ti,ab
#154 'spondylolysis'/exp
#155 'spondylolysis':ti,ab
#156 'spondylolyses':ti,ab
#157 'spondylolisthesis'/exp
#158 'spondylolisthesis':ti,ab
#159 'spondylolistheses':ti,ab
#160 'myofascial pain syndrome':ti,ab
#161 'myofascial pain syndromes':ti,ab
#162 (#116 OR #117 OR #118 OR #137 OR #138 OR #139 OR #140 OR #141 OR #142 OR #143 OR #144 OR #145 OR #146 OR #147 OR #148 OR #149 OR #150 OR #151 OR #152 OR #153 OR #154 OR #155 OR #156 OR #157 OR #158 OR #159 OR #160 OR #161)
#163 (#84 OR #116)
#164 (#27 AND #162 AND #163)
#165 [animals]/lim
#166 [humans]/lim
#167 (#165 NOT #166)
#168 (#164 NOT #167)
#169 english:la
#170 (#168 AND #169)

**Appendix 1C** CENTRAL search strategy

#1 [mh "zygapophyseal joint"]
#2 zygapophyseal:ti,ab,kw
#3 zygapophysial:ti,ab,kw
#4 facet:ti,ab,kw
#5 (#2 OR #3 OR #4)
#6 joint:ti,ab,kw
#7 joints:ti,ab,kw
#8 (#6 OR #7)
#9 (#5 AND #8)
#10 "zygapophyseal joint":ti,ab,kw
#11 "zygapophyseal joints":ti,ab,kw
#12 "zygapophysial joint":ti,ab,kw
#13 "zygapophysial joints":ti,ab,kw
#14 "facet joint":ti,ab,kw
#15 "facet joints":ti,ab,kw
#16 (#1 OR #9 OR #10 OR #11 OR #12 OR #13 OR #14 OR #15)
#17 [mh "injections, spinal"]
#18 injection:ti,ab,kw
#19 injections:ti,ab,kw
#20 intervention:ti,ab,kw

#21 interventions:ti,ab,kw
#22 (#17 OR #18 OR #19 OR #20 OR #21)
#23 (#16 AND #22)
#24 "facet joint injection":ti,ab,kw
#25 fji:ti,ab,kw
#26 facet:ti,ab,kw
#27 block:ti,ab,kw
#28 blocks:ti,ab,kw
#29 (#27 OR #28)
#30 (#26 AND #29)
#31 "facet block":ti,ab,kw
#32 "facet blocks":ti,ab,kw
#33 (#23 OR #24 OR #25 OR #30 OR #31 OR #32)
#34 [mh "back pain"]
#35 back:ti,ab,kw
#36 vertebrogenic:ti,ab,kw
#37 spine:ti,ab,kw
#38 spinal:ti,ab,kw
#39 lumbar:ti,ab,kw
#40 thoracic:ti,ab,kw
#41 (#35 OR #36 OR #37 OR #38 OR #39 OR #40)
#42 pain:ti,ab,kw
#43 pains:ti,ab,kw
#44 ache:ti,ab,kw
#45 aches:ti,ab,kw
#46 aching:ti,ab,kw
#47 (#42 OR #43 OR #44 OR #45 OR #46)
#48 (#41 AND #47)
#49 "back pain":ti,ab,kw
#50 "back pains":ti,ab,kw
#51 "back ache":ti,ab,kw
#52 backache:ti,ab,kw
#53 "back aches":ti,ab,kw
#54 backaches:ti,ab,kw
#55 "vertebrogenic pain":ti,ab,kw
#56 "vertebrogenic pains":ti,ab,kw
#57 "vertebrogenic pain syndrome":ti,ab,kw
#58 "vertebrogenic pain syndromes":ti,ab,kw
#59 dorsalgia:ti,ab,kw
#60 dorsalgias:ti,ab,kw
#61 [mh "low back pain"]
#62 "low back pain":ti,ab,kw
#63 lumbago:ti,ab,kw
#64 [mh "sciatica"]
#65 sciatica:ti,ab,kw
#66 sciatic:ti,ab,kw
#67 neuralgia*:ti,ab,kw
#68 (#66 AND #67)
#69 "sciatic neuralgia":ti,ab,kw
#70 "sciatic neuralgias":ti,ab,kw
#71 claudication:ti,ab,kw
#72 [mh "pain, referred"]

#73 refer:ti,ab,kw
#74 referred:ti,ab,kw
#75 hip:ti,ab,kw
#76 knee:ti,ab,kw
#77 leg:ti,ab,kw
#78 (#73 OR #74 OR #75 OR #76 OR #77)
#79 (#47 AND #78)
#80 "refer pain":ti,ab,kw
#81"referred pain":ti,ab,kw
#82 "hip pain":ti,ab,kw
#83 "hip pains":ti,ab,kw
#84 "knee pain":ti,ab,kw
#85 "knee pains":ti,ab,kw
#86 "leg pain":ti,ab,kw
#87 "leg pains":ti,ab,kw
#88 [mh "paresthesia"]
#89 paresthesia:ti,ab,kw
#90 paresthesias:ti,ab,kw
#91 numbness:ti,ab,kw
#92 (#48 OR #49 OR #50 OR #51 OR #52 OR #53 OR #54 OR #55 OR #56 OR #57 OR #58 OR #59 OR #60 OR #61 OR #62 OR #63 OR #64 OR #65 OR #68 OR #69 OR #70 OR #71 OR #72 OR #79 OR #80 OR #81 OR #82 OR #83 OR #84 OR #85 OR #86 OR #87 OR #88 OR #89 OR #90 OR #91)
#93 safe:ti,ab,kw
#94 safety:ti,ab,kw
#95 "side effect":ti,ab,kw
#96 "side effects":ti,ab,kw
#97 "undesirable effect":ti,ab,kw
#98 "undesirable effects":ti,ab,kw
#99 "treatment emergent":ti,ab,kw
#100 "treatment emergency":ti,ab,kw
#101 tolerability:ti,ab,kw
#102 tolerance:ti,ab,kw
#103 tolerate:ti,ab,kw
#104 toxicity:ti,ab,kw
#105 toxic:ti,ab,kw
#106 harm:ti,ab,kw
#107 harms:ti,ab,kw
#108 complication*:ti,ab,kw
#109 risk:ti,ab,kw
#110 risks:ti,ab,kw
#111 "unintended event":ti,ab,kw
#112 "unintended events":ti,ab,kw
#113 "unintended effect":ti,ab,kw
#114 "unintended effects":ti,ab,kw
#115 "adverse event":ti,ab,kw
#116 "adverse events":ti,ab,kw
#117 "adverse effect":ti,ab,kw
#118 "adverse effects":ti,ab,kw
#119 "adverse reaction":ti,ab,kw
#120 "adverse reactions":ti,ab,kw
#121 "adverse outcome":ti,ab,kw
#122 "adverse outcomes":ti,ab,kw

#123 (#93 OR #94 OR #95 OR #96 OR #97 OR #98 OR #99 OR #100 OR #101 OR #102 OR #103 OR #104 OR #105 OR #106 OR #107 OR #108 OR #109 OR #110 OR #111 OR #112 OR #113 OR #114 OR #115 OR #116 OR #117 OR #118 OR #119 OR #120 OR #121 OR #122)
#124 [mh "spinal stenosis"]
#125 [mh "spondylosis"]
#126 [mh "synovial cyst"]
#127 back:ti,ab,kw
#128 vertebral:ti,ab,kw
#129 spine:ti,ab,kw
#130 spinal:ti,ab,kw
#131 lumbar:ti,ab,kw
#132 thoracic:ti,ab,kw
#133 facet:ti,ab,kw
#134 synovial:ti,ab,kw
#135 (#127 OR #128 OR #129 OR #130 OR #131 OR #132 OR #133 OR #134)
#136 stenosis:ti,ab,kw
#137 stenoses:ti,ab,kw
#138 spondylosis:ti,ab,kw
#139 syndrome:ti,ab,kw
#140 syndromes:ti,ab,kw
#141 arthropathy:ti,ab,kw
#142 arthritis:ti,ab,kw
#143 fracture:ti,ab,kw
#144 fractures:ti,ab,kw
#145 cyst:ti,ab,kw
#146 cysts:ti,ab,kw
#147 (#136 OR #137 OR #138 OR #139 OR #140 OR #141 OR #142 OR #143 OR #144 OR #145 OR #146)
#148 (#135 AND #147)
#149 "vertebral canal stenosis":ti,ab,kw
#150 "spinal stenosis":ti,ab,kw
#151 "spinal stenoses":ti,ab,kw
#152 "thoracic spondylosis":ti,ab,kw
#153 "lumbar spondylosis":ti,ab,kw
#154 "facet syndrome":ti,ab,kw
#155 "facet joint syndrome":ti,ab,kw
#156 "facet arthropathy":ti,ab,kw
#157 "facet arthritis":ti,ab,kw
#158 "vertebral compression fracture":ti,ab,kw
#159 "spinal cyst":ti,ab,kw
#160 "facet cyst":ti,ab,kw
#161 "synovial cyst":ti,ab,kw
#162 [mh "tarlov cysts"]
#163 "tarlov cyst":ti,ab,kw
#164 "tarlov cysts":ti,ab,kw
#165 [mh "spondylolysis"]
#166 spondylolysis:ti,ab,kw
#167 spondylolyses:ti,ab,kw
#168 [mh "spondylolisthesis"]
#169 spondylolisthesis:ti,ab,kw
#170 spondylolistheses:ti,ab,kw
#171 [mh "myofascial pain syndromes"]
#172 "myofascial pain syndrome":ti,ab,kw

#173 "myofascial pain syndromes":ti,ab,kw
#174 (#124 OR #125 OR #126 OR #148 OR #149 OR #150 OR #151 OR #152 OR #153 OR #154 OR #155 OR #156 OR #157 OR #158 OR #159 OR #160 OR #161 OR #162 OR #163 OR #164 OR #165 OR #166 OR #167 OR #168 OR #169 OR #170 OR #171 OR #172 OR #173)
#175 (#92 OR #123)
#176 (#33 AND #174 AND #175)
#177 [mh animals]
#178 [mh humans]
#179 (#177 NOT #178)
#180 (#176 NOT #179)

**Appendix 1D** Scopus search strategy

(TITLE-ABS-KEY((((((zygapophyseal OR zygapophysial OR facet) AND joint*) OR "zygapophyseal joint" OR "zygapophyseal joints" OR "zygapophysial joint" OR "zygapophysial joints" OR "facet joint" OR "facet joints") AND (injection* OR intervention*)) OR "facet joint injection" OR fji OR (facet AND block*) OR "facet block" OR "facet blocks") AND ((((back OR vertebrogenic OR spine OR spinal OR lumbar OR thoracic) AND (pain* OR ache* OR aching)) OR "back pain" OR "back pains" OR "back ache" OR backache OR "back aches" OR backaches OR "vertebrogenic pain" OR "vertebrogenic pains" OR "vertebrogenic pain syndrome" OR "vertebrogenic pain syndromes" OR dorsalgia OR dorsalgias OR "low back pain" OR lumbago OR sciatica OR (sciatic AND neuralgia*) OR "sciatic neuralgia" OR "sciatic neuralgias" OR claudication OR ((refer OR referred OR hip OR knee OR leg) AND (pain* OR ache*OR aching)) OR "refer pain" OR "referred pain" OR "hip pain" OR "hip pains" OR "knee pain" OR "knee pains" OR "leg pain" OR "leg pains" OR paresthesia OR paresthesias OR numbness) OR (safe OR safety OR "side effect" OR "side effects" OR "undesirable effect" OR "undesirable effects" OR "treatment emergent" OR "treatment emergency" OR tolerability OR tolerance OR tolerate OR toxicity OR toxic OR harm* OR complication* OR risk* OR "unintended event" OR "unintended events" OR "unintended effect" OR "unintended effects" OR "adverse event" OR "adverse events" OR "adverse effect" OR "adverse effects" OR "adverse reaction" OR "adverse reactions" OR "adverse outcome" OR "adverse outcomes")) AND (((back OR vertebral OR spine OR spinal OR lumbar OR thoracic OR facet OR synovial) AND (stenosis OR stenoses OR spondylosis OR syndrome* OR arthropathy OR arthritis OR fracture* OR cyst*)) OR "vertebral canal stenosis" OR "spinal stenosis" OR "spinal stenoses" OR "thoracic spondylosis" OR "lumbar spondylosis" OR "facet syndrome" OR "facet joint syndrome" OR "facet arthropathy" OR "facet arthritis" OR "vertebral compression fracture" OR "spinal cyst" OR "facet cyst" OR "synovial cyst" OR "tarlov cyst" OR "tarlov cysts" OR spondylolysis OR spondylolyses OR spondylolisthesis OR spondylolistheses OR "myofascial pain syndrome" OR "myofascial pain syndromes")))
AND NOT (ALL(animals AND NOT humans)) AND LANGUAGE(english)

**Appendix 1E** Web of Science search strategy

(TS=((((((zygapophyseal OR zygapophysial OR facet) AND joint*) OR "zygapophyseal joint" OR "zygapophyseal joints" OR "zygapophysial joint" OR "zygapophysial joints" OR "facet joint" OR "facet joints") AND (injection* OR intervention*)) OR "facet joint injection" OR fji OR (facet AND block*) OR "facet block" OR "facet blocks") AND ((((back OR vertebrogenic OR spine OR spinal OR lumbar OR thoracic) AND (pain* OR ache* OR aching)) OR "back pain" OR "back pains" OR "back ache" OR backache OR "back aches" OR backaches OR "vertebrogenic pain" OR "vertebrogenic pains" OR "vertebrogenic pain syndrome" OR "vertebrogenic pain syndromes" OR dorsalgia OR dorsalgias OR "low back pain" OR lumbago OR sciatica OR (sciatic AND neuralgia*) OR "sciatic neuralgia" OR "sciatic neuralgias" OR claudication OR ((refer OR referred OR hip OR knee OR leg) AND (pain* OR ache*OR aching)) OR "refer pain" OR "referred pain" OR "hip pain" OR "hip pains" OR "knee pain" OR "knee pains" OR "leg pain" OR "leg pains" OR paresthesia OR paresthesias OR numbness) OR (safe OR safety OR "side effect" OR "side effects" OR "undesirable effect" OR "undesirable effects" OR "treatment emergent" OR "treatment

emergency" OR tolerability OR tolerance OR tolerate OR toxicity OR toxic OR harm* OR complication* OR risk* OR "unintended event" OR "unintended events" OR "unintended effect" OR "unintended effects" OR "adverse event" OR "adverse events" OR "adverse effect" OR "adverse effects" OR "adverse reaction" OR "adverse reactions" OR "adverse outcome" OR "adverse outcomes")) AND (((back OR vertebral OR spine OR spinal OR lumbar OR thoracic OR facet OR synovial) AND (stenosis OR stenoses OR spondylosis OR syndrome* OR arthropathy OR arthritis OR fracture* OR cyst*)) OR "vertebral canal stenosis" OR "spinal stenosis" OR "spinal stenoses" OR "thoracic spondylosis" OR "lumbar spondylosis" OR "facet syndrome" OR "facet joint syndrome" OR "facet arthropathy" OR "facet arthritis" OR "vertebral compression fracture" OR "spinal cyst" OR "facet cyst" OR "synovial cyst" OR "tarlov cyst" OR "tarlov cysts" OR spondylolysis OR spondylolyses OR spondylolisthesis OR spondylolistheses OR "myofascial pain syndrome" OR "myofascial pain syndromes")))
NOT (ALL=(animal NOT human)) AND restrict language to English
